# Evaluation of the WHO HEARTS hypertension control package in primary care clinics of rural Bangladesh: a quasi-experimental trial

**DOI:** 10.1101/2024.02.06.24302424

**Authors:** Ahmad Abrar, Jubaida Akhtar, Xiao Hu, Shamim Jubayer, Mohammad Noor Nabi Sayem, Sarmin Sultana, Mohammad Abdullah Al Mamun, Mahfuzur Rahman Bhuiyan, Fazila-tun-nesa Malik, Mohammad Robed Amin, Abdul Alim, Reena Gupta, Di Zhao, Margaret Farrell, Bolanle Banigbe, Kunihiro Matsushita, Daniel Burka, Lawrence J. Appel, Andrew E. Moran, Sohel Reza Choudhury

**Author notes:** **Corresponding Author:** Professor Sohel Reza Choudhury, Department of Epidemiology and Research, National Heart Foundation Hospital and Research Institute, Plot 7/2, Dhaka 1216, Bangladesh.

## Abstract

**Background:** The World Health Organization (WHO) promotes the HEARTS technical package for improving hypertension control worldwide, but its effectiveness has not been rigorously evaluated.

**Methods:** A matched-pair cluster quasi-experimental trial in Upazila Health Complexes (UHCs; primary healthcare facilities) was conducted in rural Bangladesh. A total of 3,935 patients (mean age 52.3 years, 70.5% female) with uncontrolled hypertension (blood pressure [BP] ≥140/90 mm Hg regardless of treatment history) were enrolled: 1,950 patients from 7 UHCs implementing HEARTS and 1,985 patients from 7 matched usual care UHCs. WHO-HEARTS package intervention components were 1) simplified treatment protocol, 2) reliable medication supply, 3) team-based care, 4) standardized follow-up, and 5) a digital information system to track patients’ BP and monitor program performance. The primary outcome was systolic BP at six months measured at the patient’s home; secondary outcomes were diastolic BP, hypertension control rate (<140/90 mm Hg), and loss to follow-up. Multivariable mixed-effect linear and Poisson models were conducted as appropriate.

**Results:** Baseline mean systolic BP was 158.4 mm Hg in the intervention group and 158.8 mm Hg in the usual care group. At six months, the primary outcome was obtained in 95.5% of participants. Compared to usual care, the intervention significantly lowered systolic BP (-23.7 mm Hg vs. -20.0 mm Hg; net difference -3.7 mm Hg, p<0.001) and diastolic BP (-10.2 mm Hg vs. -8.3 mm Hg; net difference -1.9 mm Hg, p<0.001) and improved hypertension control (62.0% vs. 49.7%, net difference 12.3%, p<0.001). The occurrence of missed clinic visits was lower in the intervention group (8.8% vs. 39.3%, p<0.001).

**Conclusions:** In rural Bangladesh, WHO-HEARTS package implementation significantly lowered BP and improved hypertension control. These results support scale up of the WHO-HEARTS hypertension control package in Bangladesh and its implementation in other low- and middle-income countries. [clinicaltrials.gov registration ID NCT04992039]

**Clinical Perspective:**

- The Global Hearts Initiative is implementing a standard World Health Organization (WHO) HEARTS package for hypertension control in primary care clinics of 32 low- and middle-income settings. This quasi-experimental trial was completed alongside HEARTS program expansion in rural Bangladesh and is the first to rigorously assess the complete HEARTS package for hypertension.
- Compared with usual care, the WHO-HEARTS package significantly lowered blood pressure and improved hypertension control in hypertensive patients.
- WHO-HEARTS package implementation was feasible and effectively improved hypertension control in rural Bangladesh. The WHO-HEARTS is a standard and effective approach to improving hypertension control in low- and middle-income countries.

## Introduction

High blood pressure (BP) is the leading preventable cause of death worldwide, accounting for over 10 million annual preventable deaths. Four out of every five hypertension patients live in low- and middle-income countries (LMICs).^1^ According to the 2018 National STEPS Survey of NCD Risk Factors in Bangladesh, of the estimated 21 million adults with hypertension, 54% were unaware of their hypertension, 18% were diagnosed but untreated, 17% were treated but uncontrolled, and only 11% had BP controlled <140/90 mm Hg.^2^

To improve hypertension control worldwide, the World Health Organization (WHO) in 2016 developed the HEARTS technical package,^3^ a practical, public health approach to scaling up national hypertension control programs that is consistent with WHO hypertension treatment guidelines.^4^ HEARTS recommends 1) simple treatment protocols with specific antihypertensive medications and doses, 2) strategies to ensure a reliable supply of affordable, quality-assured antihypertensive medications, 3) team-based care including non-physician health workers, 4) a patient-centered approach including community-based hypertension care services, and 5) a robust health information system for tracking hypertension patients and program performance over time. By 2022, over 12 million patients were enrolled in HEARTS hypertension treatment programs in 32 countries,^5^ yet the complete HEARTS package has never been rigorously evaluated.To increase hypertension detection and control in Bangladesh, the Bangladesh Ministry of Health and Family Welfare (MOHFW) and the National Heart Foundation of Bangladesh (NHF-B) in collaboration with Resolve to Save Lives (RTSL) developed and launched the Bangladesh Hypertension Control Initiative (BHCI), a HEARTS-based hypertension control program in four health care facilities in 2018. In the context of an expansion in 2020 of the HEARTS program to new health care facilities in rural Bangladesh, this study tested the effects of the WHO HEARTS package implementation on hypertension outcomes.

## Methods

### Study design

The Bangladesh HEARTS Trial, a matched-pair cluster quasi-experimental trial, was conducted in 14 primary healthcare facilities named Upazila Health Complexes (UHCs) in rural Bangladesh to assess the effectiveness of the WHO HEARTS package. Seven UHCs that had planned to implement the HEARTS package under ongoing expansion of the BHCI program were selected as intervention sites. Six UHCs in the geographically separated Jamalpur district plus one geographically similar UHC in Habiganj district served as the usual care (control) UHCs. The seven intervention sites and the seven usual care sites were matched on population size and literacy. Geographically isolated UHCs were purposely selected to be intervention sites to reduce the risk of cross-facility contamination.

### Study participants

Inclusion criteria were age 18 years or older, diagnosis of hypertension (known prior diagnosis or a new diagnosis), and a baseline systolic BP ≥140 mm Hg or diastolic BP ≥90 mm Hg (i.e., “uncontrolled hypertension”) regardless of whether the person was taking antihypertensive medication. Exclusion criteria were current pregnancy, current treatment of an acute illness, terminal illness, or unwillingness to provide informed consent.

### Recruitment

An identical opportunistic hypertension screening protocol was implemented in intervention and usual care sites. All patients aged 18 years and older who presented to the outpatient reception desk of the UHC for a non-emergency visit had their BP measured with an “arm in” automated oscillometric digital BP device (A&D TM 2657P). For patients found to have systolic BP ≥130 or diastolic BP ≥80 mm Hg on the first measurement, a second BP was measured by a nurse after an interval of two minutes. If the second BP was systolic ≥140 or diastolic ≥90 mm Hg, the patient was referred to a medical officer on the same day for a third, confirmatory BP. A hypertension diagnosis was made if the third, confirmatory BP by a UHC medical officer was systolic ≥140 or diastolic ≥90 mm Hg. In some instances, due to staffing shortages, trained field research staff measured 2^nd^ and 3^rd^ BP in place of a UHC nurse and Medical Officer; this was occasionally required in both intervention and usual care UHCs. Both the 2^nd^ and 3^rd^ BP measurements were performed using a validated desktop oscillometric digital BP measurement device (Omron Model HBP-1120). Patients diagnosed with HTN were provided information about the HEARTS trial, and for those agreeing to participate in the trial, informed consent was obtained. All eligible patients were invited to enroll, irrespective of their antihypertensive medication use at baseline. Age, sex, history of comorbidities, prior hypertension treatment were self-reported at baseline.

### Hypertension management

Consented patients with confirmed hypertension at the intervention sites received hypertension care according to the WHO HEARTS technical package. As shown in **Supplementary Table 1**, key components of the HEARTS intervention were 1) use of a three-step, drug-and dose-specific hypertension treatment protocol (**Figure 1**), 2) standardized inventory and procurement practices to ensure a reliable supply of protocol medications, 3) team-based care involving Medical Officers (physicians), nurses and medical assistants, 4) a standardized approach to follow-up and retain patients in care, and 5) a digital health information system (the Simple mobile application, www.simple.org) for registering patients into a secure database and tracking patient hypertension management and program performance over time.^6^ Intervention UHC patients who missed a scheduled follow-up visit were contacted by phone and advised to attend the UHC at earliest convenience.

**Figure 1:**
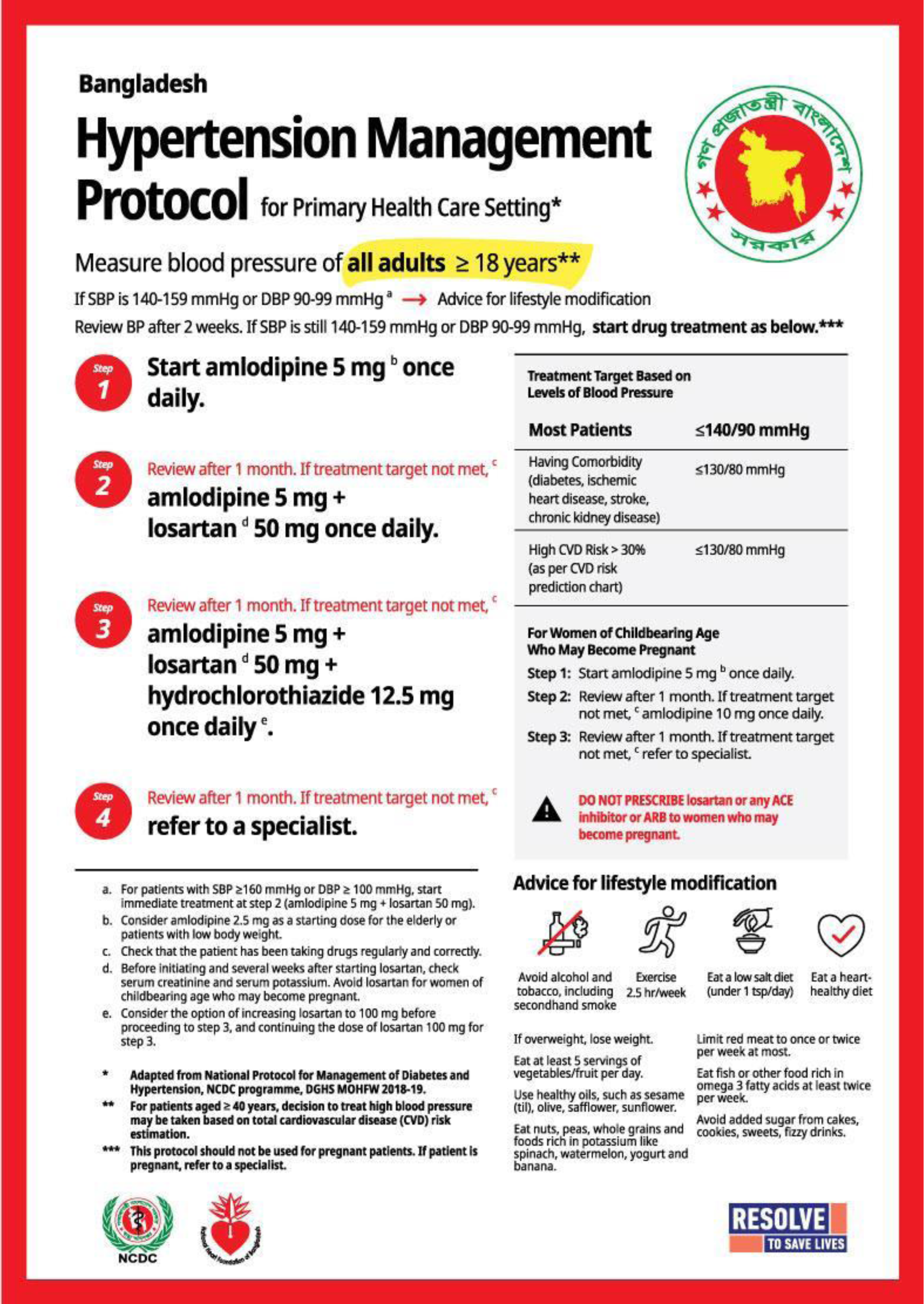
Drug and dose specific hypertension management protocol used in the Bangladesh HEARTS Hypertension Control Initiative.

Usual care site patients received the less structured hypertension management services commonly provided in UHCs outside of the HEARTS program (**Supplementary Table 1**). Specifically, there was no standardized treatment protocol, no expanded team-based care, no schedule of systematic patient follow-up, and there were no added efforts to improve adherence and retention in care.

There were some features similar in both the intervention and usual care sites. Nurses were trained on BP measurement and to use the Simple app to record BP measurements and medication data. Both intervention and usual care site patients had access to hypertension medications free of charge.

### Endline blood pressure outcomes

The primary outcome was the difference between the intervention and usual care sites in change of systolic BP from the baseline measurement to a follow up measurement six months later. Because of severe flooding at several sites at approximately six months, follow-up was extended in both groups by two months, if needed, in order to complete the follow-up visit to assess endline BP. The mean length of observation time (±standard deviation) from baseline to endline measurement was 238.9±26.4 days in intervention group and 233.5±29.0 days in usual care group. The research team decided to obtain endline BP in homes, instead of the UHC clinics, to obtain high ascertainment of trial outcomes and minimize the potential impact of differential loss-to-follow-up in the usual care and intervention sites. At the six month follow-up visit, research staff obtained triplicate measures of BP in the participants’ homes in both the intervention and usual care clusters using standard technique and the same Omron Model HBP-1120 used during the baseline assessment. The average of the 2^nd^ and 3^rd^ readings represented each participant’s endline BP.

Secondary outcomes were between-group difference in change of diastolic BP, hypertension control rate at six months, loss to follow-up, and early follow-up rate. Hypertension control rate was defined as the number of enrolled patients with controlled BP (systolic BP <140 and diastolic BP <90 mm Hg) measured in the community at six months divided by the number of enrolled patients who completed the 6-month follow up visit. Loss to follow-up rate was defined as the number of enrolled patients with no clinic visit through the entire 6-month follow up period divided by the number of enrolled patients who completed the 6-month follow up visit. Early follow-up rate was defined as enrolled patients with clinic visit within the first 3 months after enrollment divided by the number of enrolled patients who completed the 6-month follow up visit.

### Endline questionnaire

Along with measuring the endline BP, patients were also invited to provide verbal responses to a standard questionnaire (**Supplemental Material appendix 1**). The endline questionnaire included medication information including dose and dosage frequency, medication adherence for the last 7 days, experience of medicine-related adverse effects, incident hypertension-related complications or hospitalizations, and other implementation outcomes. Questionnaire responses assessed between-group differences in implementation outcomes between the HEARTS intervention and usual care groups. In response to the questions related to level of satisfaction with the quality of hypertension care received at the UHC in the past six months, those who responded ‘satisfied’ or ‘very satisfied’ were categorized as satisfied. Responses indicating ’agree’ or ’strongly agree’ to the question on self-perceived improvement in hypertension self-management were classified as ’improved ability’. Responses from participants expressing ‘definitely yes’ or a ‘possibly’ about the intention to visit UHC again for continued hypertension treatment were classified as ’again’. Participants who received treatment for hypertension elsewhere before visiting the study UHC were asked to compare their experience about out-of-pocket expenditure for hypertension treatment. A response with ‘yes’ was considered as spending ‘less money’ since coming to UHC for hypertension treatment. Participants reporting they were unable to pay any medical bills in past 12 months were defined as having had a ‘bill problem’. A medication intensity score was calculated by standardizing use of multiple antihypertensive drugs with different doses and dosage frequencies to assess the between-group difference in use of medication at endline. The endline questionnaire collected names, dosage and frequency of 17 antihypertensive medicines. To calculate the medication intensity score for one medicine, the prescribed daily dose for this medicine was set as the numerator. The denominator was the daily standard dose, as sourced from UpToDate (uptodate.com) and MayoClinic.org. For example, the standard dose for amlodipine is 5 mg (Supplementary Table 1). If a participant was taking amlodipine once a day with a 10 mg dosage each time, the score for amlodipine would be 10 mg*1/5 mg =2. If a medicine listed was not used by a participant, the score for this medicine was simply 0. This score was calculated for each individual medicine and summed up for all of the patient’s medicines to derive a total score, used as the medication intensity score for that participant. The score was computed for each participant who completed the endline questionnaire. Adherence to the medication was assessed by self-reported missing one or more days of medication in the week prior to the endline follow-up visit. Given the unexpected flooding, in the questionnaire, participants were also asked whether flooding prevented them from visiting the UHC or getting a refill of medication in the past 2 months.

### Statistical analysis

A sample size of 2,100 was estimated to provide a minimal detectable difference of 5 mm Hg in systolic BP between the intervention and usual care groups with 150 participants in each cluster, assuming a type I error of 5%, power of 80%, mean (SD) systolic BP of 148 (20) mm Hg at baseline, intra-cluster correlation coefficient of 0.01,^7^ follow-up rate of 50% (based on experience in an initial pilot), and a coefficient of variation of cluster size of 0.65.^8^ This sample size was planned for enrollment over a three-month period. Observing an overwhelming response from patients seeking to be enrolled into hypertension care at both HEARTS intervention and usual care sites, researchers decided to continue the enrollment for three months as planned even after the initial sample size goal was reached. The final enrollment number was 3,935.

Descriptive statistics summarized baseline characteristics of the intervention and usual care sites. To evaluate the effect of intervention on the changes of systolic and diastolic BP over follow-up, we used linear mixed effect models with a three-level hierarchical approach. The first level represents within-person variation with multiple BP measurements over time. The second level represents the variation of BP across participants, and the third level represents the variation across clusters. Between group differences (95% confidence intervals [CI]) in BP change over six months were evaluated from the interaction term between the intervention group and the visit (follow-up vs baseline). Given the small number of clusters, we adopted unmatched analysis as the main statistical analytical method.^9^

We also evaluated the effect of intervention on hypertension control rate, follow-up rate and patient-reported features of care, including the quality of hypertension care, reported improved BP management, planning to visit UHC again to receive ongoing treatment, spending less money coming to UHC compared to seeking treatment elsewhere, and having billing problems. We used mixed-effects Poisson regression with robust variance to estimate the incidence rate ratio (IRR) and 95% CI for these outcomes comparing intervention vs. usual care group. Additionally, we calculated the marginally adjusted rate of these outcomes for each group and the mean differences.

For all analyses, we used a intention-to-treat analysis approach and adjusted for individual-level confounders including intervention group, age, sex, baseline assessment of history of heart attack, stroke, chronic kidney disease, diabetes, prior hypertension treatment, whether a severe flooding event that occurred at about six month’s follow up affected medication refill, as well as cluster-level confounders including population size, area size, and literacy rate. For outcomes of BP changes over time, we also incorporated interaction terms between visit and these baseline individual-level covariates.

We conducted subgroup analyses in strata defined by age, sex, baseline BP, hypertension medication use, and diabetes. For sensitivity analyses, we used two-level mixed effect models at the individual to estimate the differences in BP change within each matched cluster pair, then pooled the estimates from all cluster pairs together using meta-analysis random-effect model.

### Ethical considerations

IRB approval was obtained from the National Ethics Review Committee of the Bangladesh Medical Research Council (BMRC) and the internal review board of Vital Strategies (the organization that Resolve to Save Lives was part of at that time). This trial was registered at ClinicalTrials.gov. (Trial registration ID NCT04992039). All participants provided written informed consent.

## Results

### Participant enrollment at baseline and follow-up at six months

Among 9,056 adults screened for eligibility (5,692 in intervention group, 3,364 in usual care), 5,121 participants did not meet the inclusion criteria (3,742 in intervention group, 1,379 in usual care) (**Figure 2**). The study population therefore included 3,935 participants (1,950 in intervention group, 1,985 in usual care group). The number of patients lost-to-follow up at the time of the six-month endline visit was 47 (2.4% of enrolled) in intervention group and 81 (4.1% of enrolled) in usual care group. Twelve patients had discontinued hypertension treatment at UHCs (5 in intervention group and 7 in usual care group). There were 37 deaths during follow up (16 in intervention group and 21 in usual care group). Finally, 3,758 participants (95.5%) remained in the analysis [1882 in intervention group (96.5%), 1876 in usual care group (94.5%) ] after excluding those who were not available for the follow-up at the six month endline visit for above reasons.

**Figure 2:**
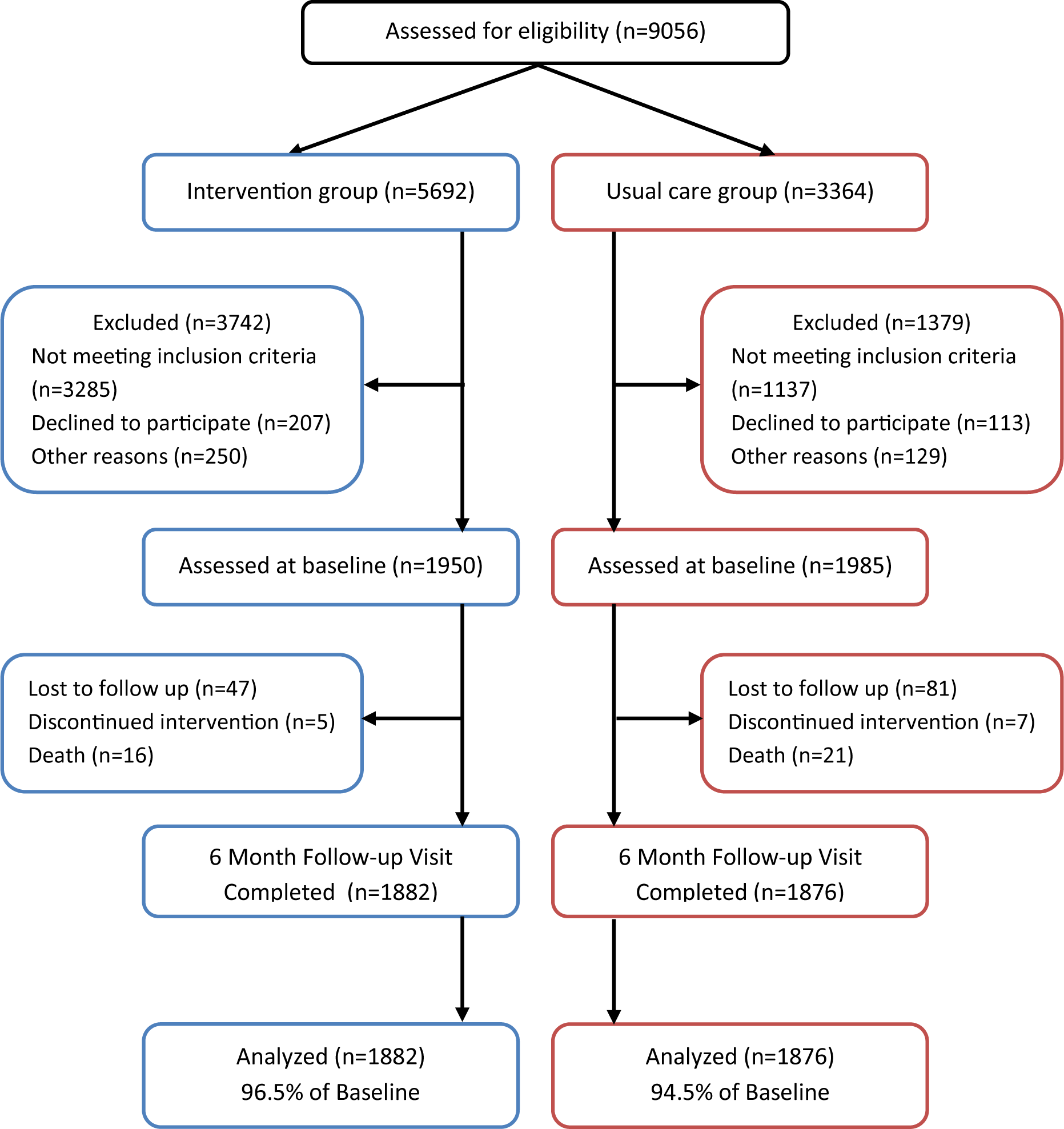
Inclusion and exclusion flowchart, the Bangladesh HEARTS trial.

### Participant characteristics

At baseline, mean (±SD) age of the participants was 52.3±12.4 years, and 70.5% were female; less than 1% had a history of heart attack, stroke, or chronic kidney disease. Baseline characteristics were generally similar in the intervention and usual care groups; however, the intervention group had higher use of antihypertensive medication at baseline (92.2% vs 68.8%) and a higher prevalence of diabetes (19.4% vs 7.5%; **Table 1**).

**Table 1:** Baseline characteristics, the Bangladesh HEARTS trial*.

|  | Total | Intervention | Usual Care | p-value |
| --- | --- | --- | --- | --- |
|  | N=3,935 | N=1,950 | N=1,985 |  |
| Female | 2,775 (70.5) | 1,350 (69.2) | 1,425 (71.8) | 0.079 |
| Age (years) | 52.3 (12.4) | 53.3 (12.2) | 51.3 (12.5) | <0.001 |
| Diagnosis of diabetes | 526 (13.4) | 378 (19.4) | 148 (7.5) | <0.001 |
| Prior heart attack | 20 (0.5) | 10 (0.5) | 10 (0.5) | 0.97 |
| Prior stroke | 36 (0.9) | 19 (1.0) | 17 (0.9) | 0.70 |
| Prior CKD | 6 (0.2) | 3 (0.2) | 3 (0.2) | 0.98 |
| Baseline HTN medication use | 3,164 (80.4) | 1,798 (92.2) | 1,366 (68.8) | <0.001 |
| Baseline SBP (mm Hg) | 158.6 (15.2) | 158.4 (15.0) | 158.8 (15.4) | 0.48 |
| Baseline DBP (mm Hg) | 92.3 (10.3) | 92.3 (10.2) | 92.2 (10.3) | 0.86 |

\*Data are presented as mean (SD) for continuous measures, and n (%) for categorical measures.
Abbreviations: SBP- systolic blood pressure; DBP- diastolic blood pressure, HTN- hypertension, CKD- chronic kidney disease.
Those with “unknown” for prior disease history were assumed to have no relevant disease history.

### BP change and hypertension control at six months

At the six-month follow-up visit, mean adjusted systolic BP was 23.7 (95% CI, 22.7 to 24.7) mm Hg lower than baseline in the intervention group and by 20.0 (95% CI, 19.1 to 21.0) mm Hg lower than baseline in the usual care group. The intervention group experienced a greater systolic BP reduction than the usual care group (net adjusted difference of -3.7 (95% CI, -2.2 to -5.1) mm Hg; **Figure 3a**). Mean adjusted diastolic BP was reduced by 10.2 (95% CI, 9.7 to 10.8) mm Hg in the intervention group and by 8.3 (95% CI, 7.8 to 8.9) mm Hg in the usual care group. The intervention group experienced a greater diastolic BP reduction than the usual care group (net adjusted difference of -1.9 (95% CI, -1.1 to -2.7) mm Hg; **Figure 3b**). At six months, hypertension control was 62.0% (95% CI, 59.3 to 64.8%) in the intervention group and 49.7% (95% CI, 46.7 to 52.6%) in the usual care group (net adjusted difference of 12.4% (95% CI, 8.0 to 16.8%); **Figure 3c**). In comparison with primary adjusted analyses, results of the crude analyses showed slightly lesser differences in BP and hypertension control (**Supplemental figures 2a, 2b, and 2c**).

**Figure 3a:**
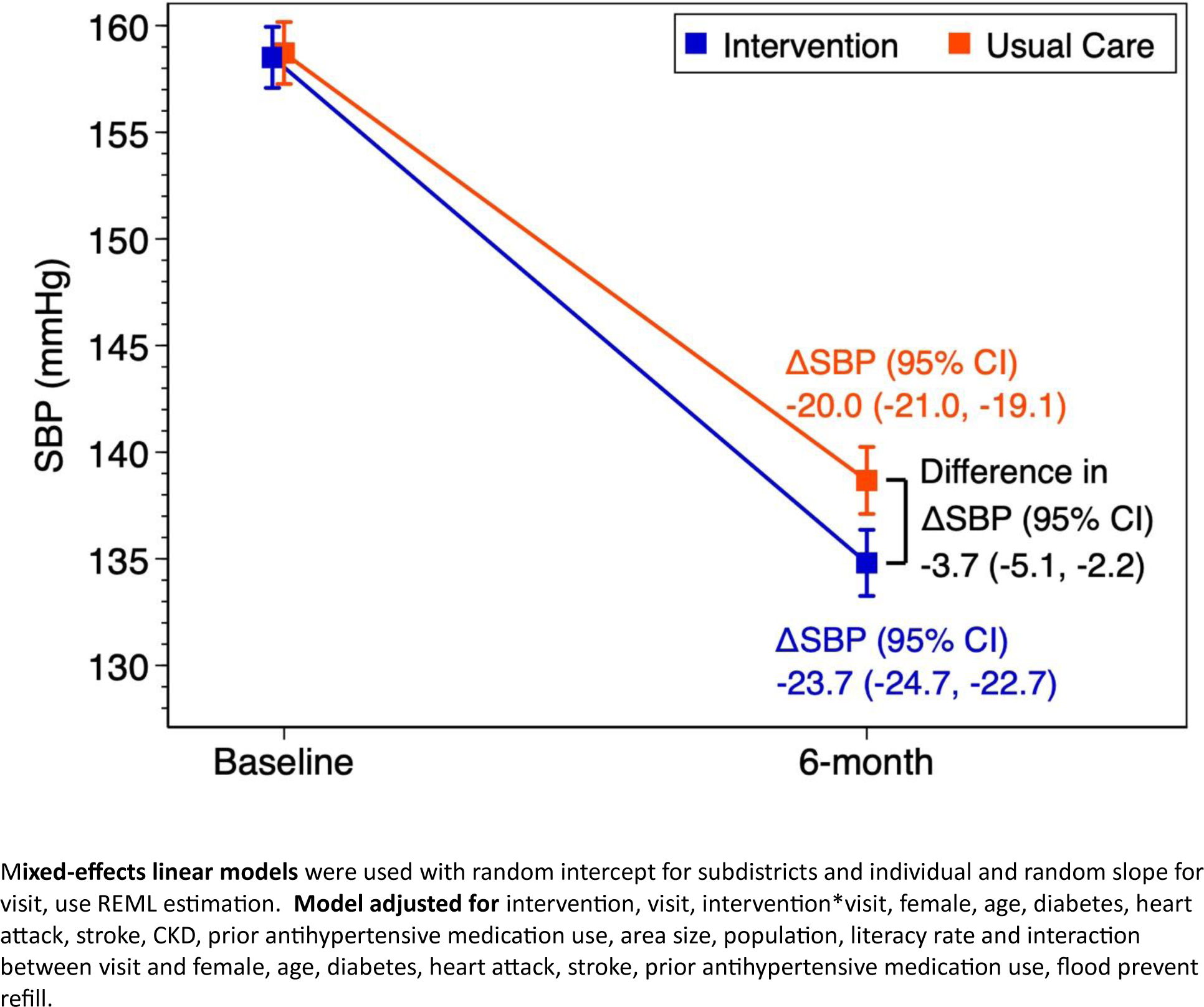
Adjusted difference in mean baseline and endline systolic blood pressure and changes at six months, the Bangladesh HEARTS trial.

**Figure 3b:**
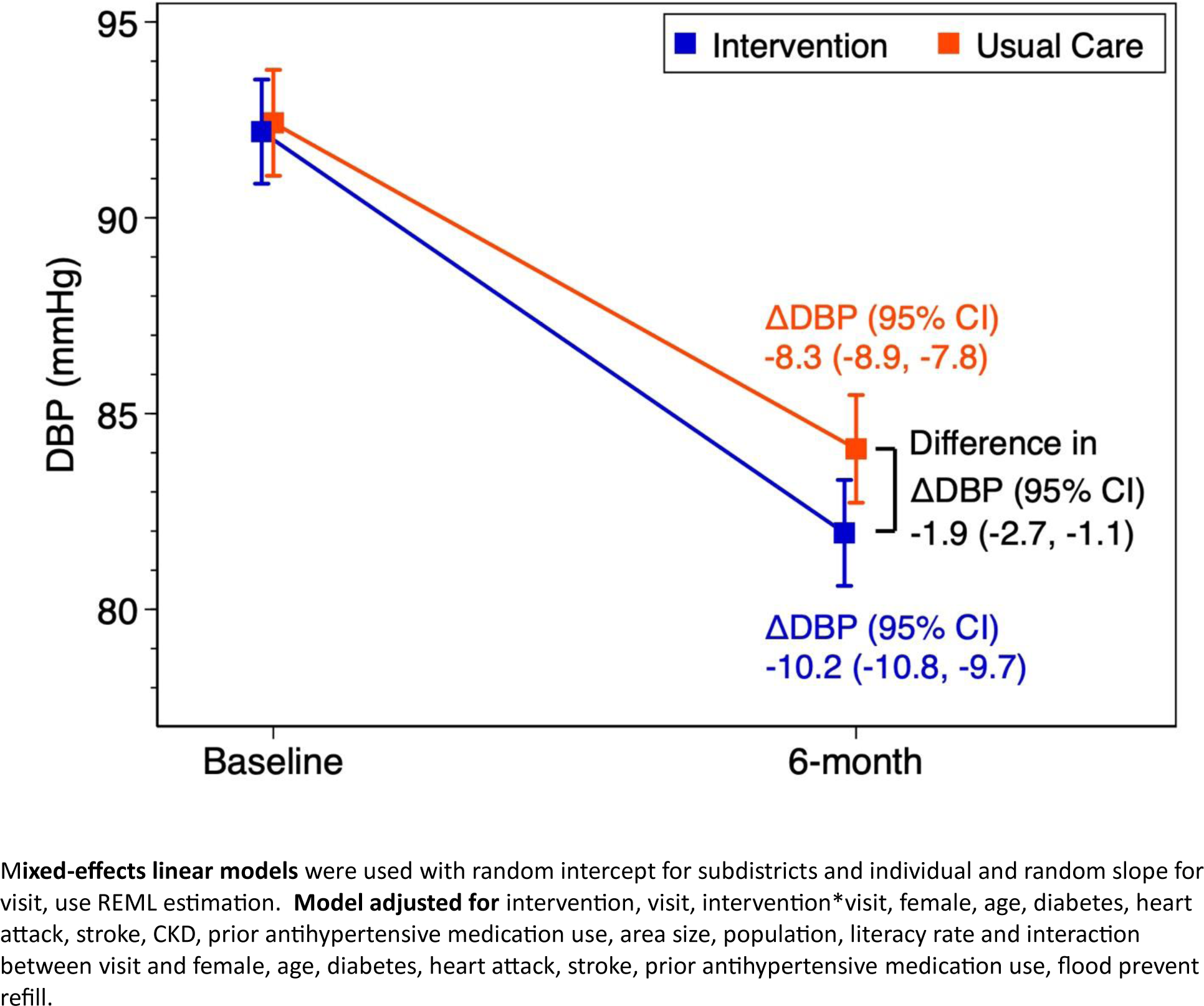
Adjusted difference in mean baseline and endline diastolic blood pressure and changes at six months, the Bangladesh HEARTS trial.

**Figure 3c:**
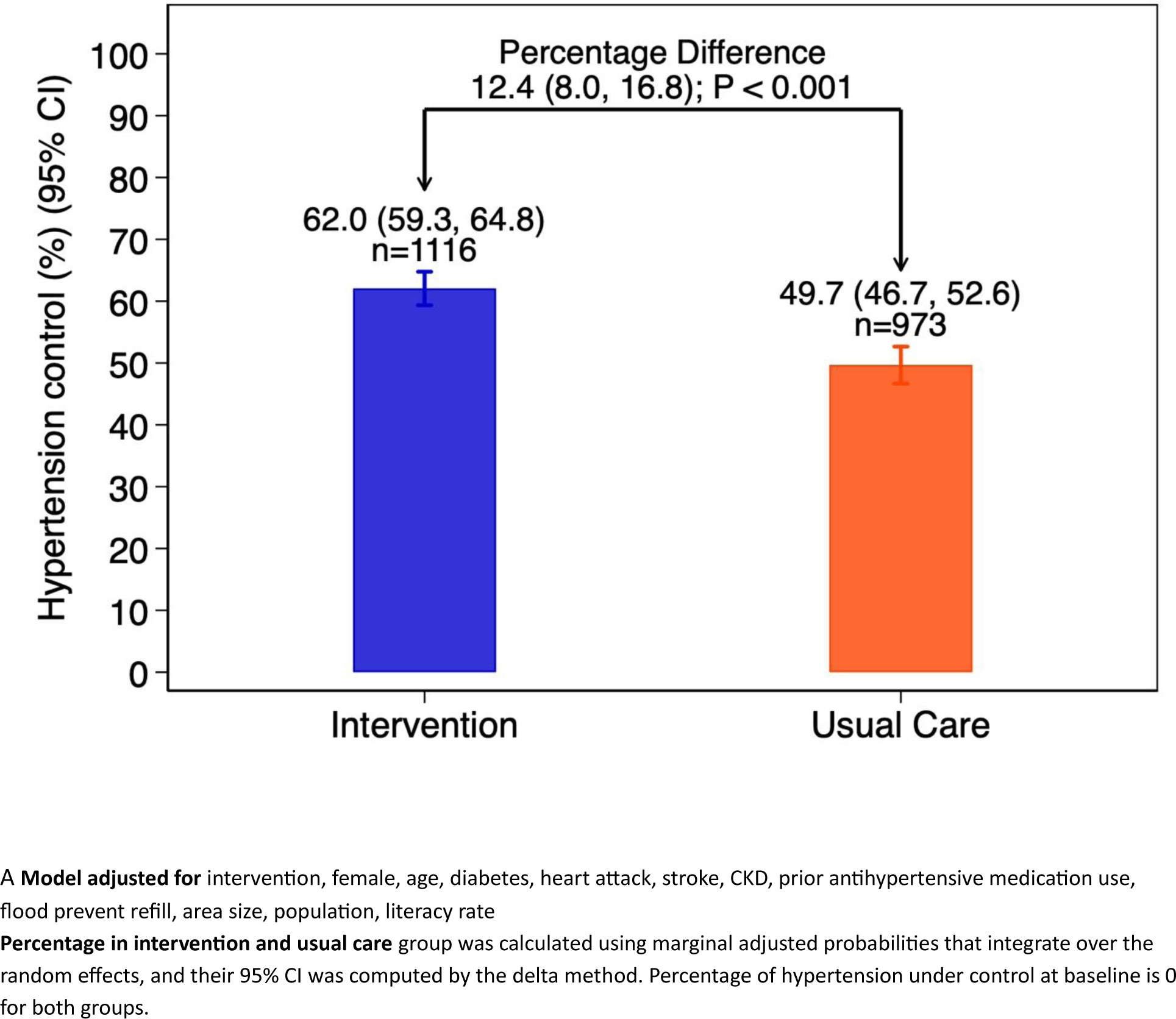
Adjusted difference in hypertension control at six months.

### Subgroup analyses

In pre-specified subgroup analyses of systolic BP, diastolic BP, and hypertension control (**Figures 4a, 4b, and 4c**), the effect of the intervention group was always greater than that of the usual care group; in several instances, the subgroup difference in between-group outcomes was statistically significant. The subgroups that appeared to have greater benefits from the intervention group were persons older than age 55 years, men, and those not on medication at baseline.

**Figure 4:**
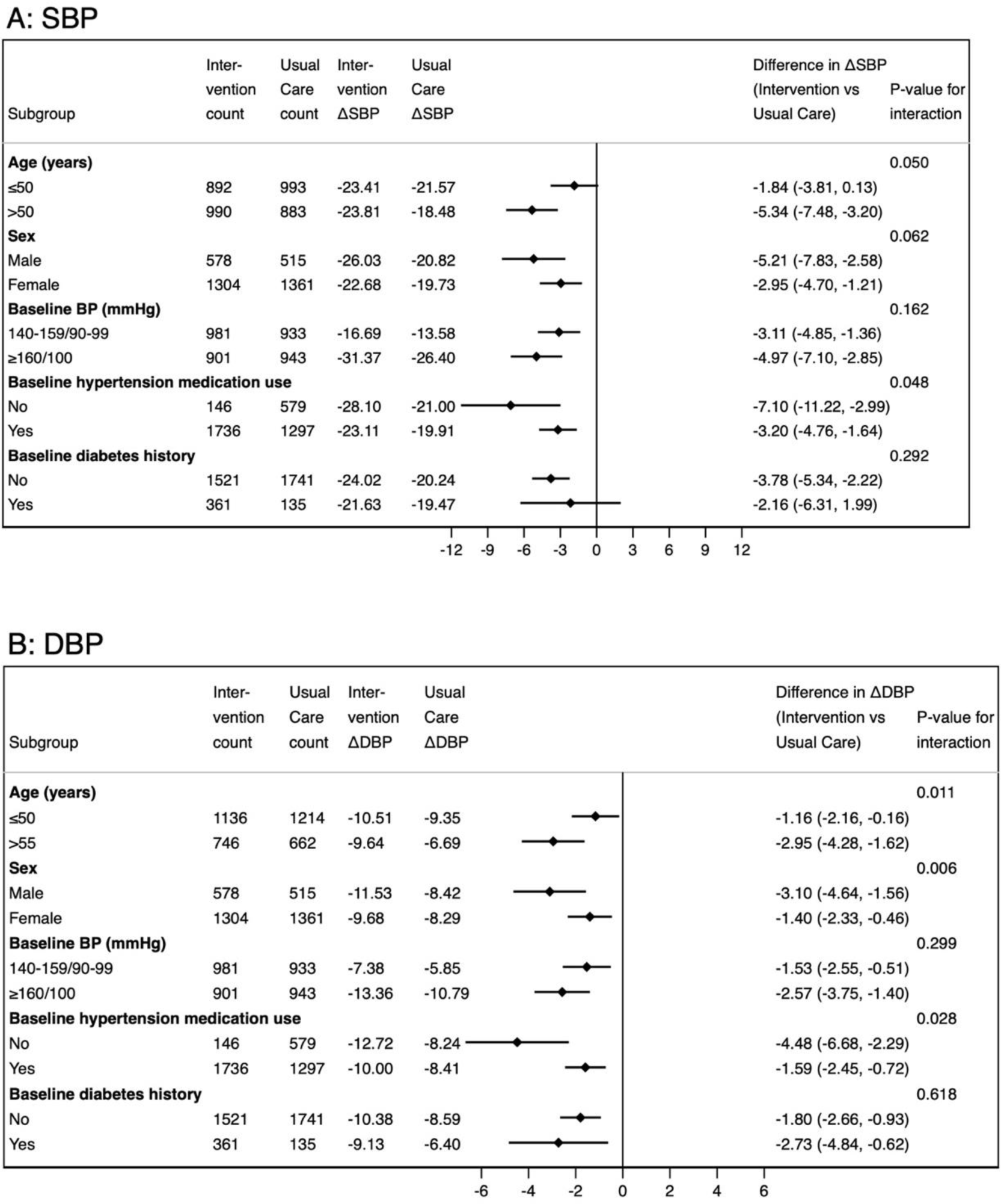
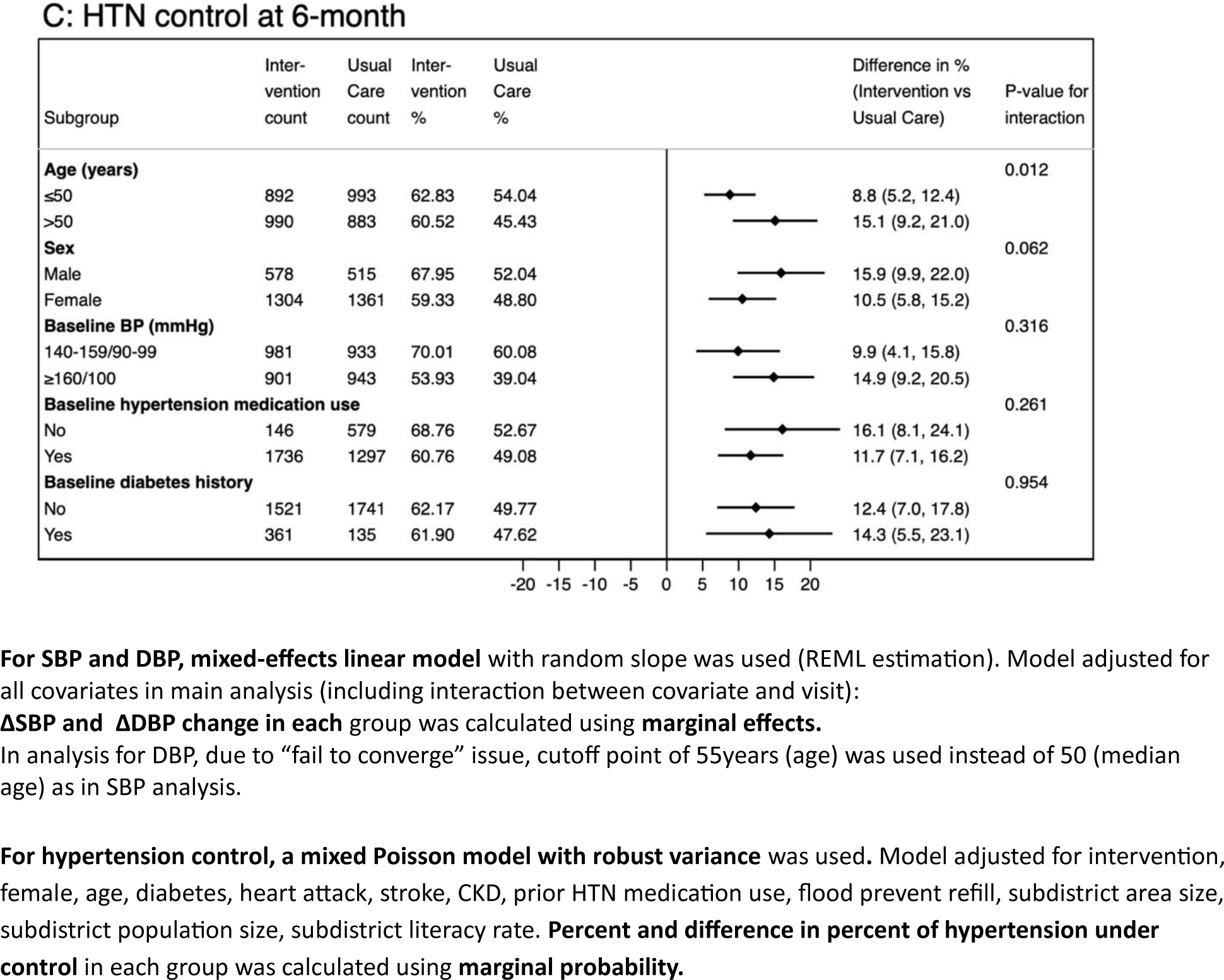
Subgroup analysis results for between group differences in change in systolic BP (4A), diastolic BP (4B), and proportion with hypertension control (4C) at the six month follow up visit, the Bangladesh HEARTS trial.

### Implementation outcomes at six months

Compared to patients in the usual care group **(Table 2)**, patients in the HEARTS intervention group were less likely to miss scheduled clinic visits (incidence rate ratio [IRR] 0.22, 95% CI: 0.15 to 0.32), and more likely to have a clinic visit within the first 3 months after enrollment (IRR 1.67, 95% CI: 1.49 to 1.87). The medication intensity score at endline, standardized for dose and frequency, was higher in the intervention group patients compared to the patients in the usual care group (1.52, 95% CI: 1.48 to 1.56 vs. 1.01, 95% CI: 0.97 to 1.05). (**Supplementary Table 4**)

**Table 2:** Incidence rate ratio (IRR) for intervention and probability difference comparing intervention and control arm.

|  | Lost to follow up | Early follow up | Satisfied | Improved | Again | Less money | Bill problem |
| --- | --- | --- | --- | --- | --- | --- | --- |
| **IRR for Intervention** | **0.22**<br>**(0.15, 0.32)** | **1.67**<br>**(1.49, 1.87)** | **1.13**<br>**(1.05, 1.22)** | **1.25**<br>**(1.05, 1.49)** | 1.03<br>(0.99, 1.07) | **1.36**<br>**(1.19, 1.56)** | 1.00<br>(0.76, 1.32) |
| **Count** |  |  |  |  |  |  |  |
| Intervention | 150 | 1667 | 1731 | 1563 | 1773 | 1296 | 931 |
| Control | 802 | 919 | 1526 | 1188 | 1706 | 754 | 812 |
| **Marginal probability (%)** |  |  |  |  |  |  |  |
| Intervention | 8.8 (6.1, 11.4) | 85.3<br>(82.2, 88.3) | 91.9<br>(90.3, 93.6) | 81.2<br>(77.6, 84.8) | 94.0<br>(92.8, 95.1) | 88.6<br>(84.3, 92.8) | 46.5<br>(41.9, 51.1) |
| Control | 39.3<br>(33.9, 44.7) | 50.9<br>(45.5, 56.3) | 81.4<br>(75.3, 87.5) | 64.8<br>(54.8, 74.8) | 91.2<br>(87.8, 94.6) | 65.0<br>(57.4, 72.5) | 46.4<br>(34.2, 58.6) |
| Difference | **-30.5 (-37.3, -23.7)** | **34.3 (28.0, 40.6)** | **10.5 (4.3, 16.8)** | **16.4 (4.8, 28.1)** | 2.8 (-0.8, 6.4) | **23.6 (14.3, 32.8)** | 0.1 (-12.5, 12.8) |

**Mixed Poisson model with robust variance** was used. Model adjusted for intervention, female, age, diabetes, heart attack, stroke, CKD, prior HTN medication use, flood prevent refill, subdistrict area size, subdistrict population size, subdistrict literacy rate. **Percent and difference in percent** were calculated using **marginal probability**. Bold font indicates statistical significance (p-value <0.05).

Patients in the intervention group were more satisfied with the quality of hypertension care received at the HEARTS program UHCs (91.9% vs. 81.4%, p=0.002), felt they had improved their ability to manage their hypertension (81.2% vs. 64.8%, p=0.011), and spent less money for their hypertension care compared with usual care patients (88.6% vs 65.0%, p<0.001) (**Table 2**). Patients in the intervention group had higher self-reported medication adherence and were more likely to be taking hypertension medications at follow-up (i.e. taking antihypertensive medication at endline 95.4% in the intervention group and 75.7% in usual care group, p<0.001, and missing at least 1 day of medication in the last week was 41.2% in the intervention group and 99.9% in the usual care group, p<0.001) (supplementary table 7).

During the study, there was an unexpected severe flooding event that impacted both intervention and usual care groups, which occurred towards the end of follow-up. The intervention group was more affected by the flooding than the usual care group. Specifically, compared to patients in the usual care group, those in the intervention group reported that flood prevented them to attend one or more clinic visits (30% vs 8%, P<0.001) and flood prevented them to receive at least one medication refill (27% vs 9%, P<0.001).

## Discussion

In this matched-pair cluster, quasi-experimental trial conducted in primary healthcare facilities in rural Bangladesh, we documented that implementation of the WHO-HEARTS hypertension control package significantly lowered both systolic and diastolic BP, and improved hypertension control compared with usual care. The HEARTS package appeared effective in all pre-specified groups, with some evidence of greater benefits in older persons, men, those with newly diagnosed hypertension, and those not taking anti-hypertensive medication at baseline.

The outcomes of the Bangladesh HEARTS trial align with other pragmatic trials of primary care-based trials based in LMIC settings testing a public health approach to hypertension control. Although none of these studies specifically tested the WHO-HEARTS package, they often included features similar to WHO-HEARTS, e.g, improved access to medicines, team-based, patient-centered approach, and systematic follow up).^10–13^ In these earlier trials, conducted in Bangladesh/Pakistan/Sri Lanka, Argentina, Ghana, and China, the systolic BP change difference between intervention and usual care/control varied between 3.6 and 14.5 mm Hg. Difference in hypertension control at endline varied between 5.2 and 32.5 percentage points. Notably, the Bangladesh HEARTS trial observed outcomes only at 6 months, whereas the earlier pragmatic hypertension control trials observed outcomes from between 12 and 24 months, and most did not observe substantial intervention benefits until after 6 months of intervention effect. The prior trial reporting the largest effect size, the China Rural Hypertension Control Project trial, 90% of intervention participants received free or discount-priced medicines, but not the usual care group had higher out-of-pocket medicine costs, whereas in Bangladesh HEARTS and the other prior trials, participant hypertension medicine costs were similar in intervention and usual care groups.

Process measures gathered at six months suggested superior HTN control program performance at HEARTS intervention sites compared with usual care sites. In fact, we observed that the HEARTS program demonstrated improvements in several primary drivers of improved BP control, including higher retention in care and clinic visit frequency, medication adherence, and medicaton treatment intensity score. Visit frequency, treatment intensity, and medication adherence are strongly correlated with improved blood pressure control and likely contributed to the higher hypertension control observed in the intervention sites.^14,15^ A prior costing study established the budget impact of Bangladesh HEARTS program implementation.^16^ Nonetheless, even at the intervention sites there is ample room for continous quality improvement in these processes for the Bangladesh HEARTS program.

The endline survey also suggested superior patient-centered outcomes in the HEARTS program compared with usual care,. Patients in the intervention group reported greater ability to manage their hypertension, higher satisfaction with the quality of care provided from the primary healthcare facilities of rural Bangladesh, and reduced out-of-pocket expenditures for hypertension care. Though between group differences in implementation outcomes were modest, the overall pattern of results was uniformly in favor of the HEARTS intervention.

Limitations of the study include a quasi-experimental design, rather than a randomized trial design. As a result of study design, there were some imbalances in baseline characteristics, with greater number of ‘’hard-to-control” patients (e.g., those with diabetes and uncontrolled BP despite baseline anti-hypertensive medications) in the intervention group compared to the control group. Indeed, results were even strengthened after adjustment for multiple baseline covariates. Interestingly, subgroup analysis showed a much greater systolic BP reduction of -7.5 mm Hg between intervention and usual care participants not on medications at baseline. Second, unexpected severe flooding interrupted delivery of care and medication refills, more so in the intervention sites. Hence, it is possible that without the flooding, there would have been even greater benefits in the intervention group. Third, the follow-up period was relatively short, just six months; nonetheless, the vast majority of patients in the intervention group received treatment and achieved a high level of hypertension control, i.e., 62%.

Fourth, the study had different types of BP measurements at baseline (clinic measurements) and at 6 months (home measurements). Home measurements were selected because of concern that many patients would not have a clinic visit at six months and that there would be a substantial difference between usual care and intervention sites with the attendant risk of biased ascertainment. As displayed in Table 2, this imbalance in clinic attendance did occur. In this context, the home BP, obtained in the vast majority of participants (>95%) in both intervention and usual care sites, was a more unbiased assessment of trial outcomes (difference between groups); however, use of home-measured BP at six months might overestimate the magnitude of change from baseline.

Strengths of this study include enrollment of a diverse cohort of persons with uncontrolled hypertension in rural Bangladesh, evaluation of a standardized package of hypertension control services package recommended by WHO and endorsed by Bangladesh’s national government, high retention in clinical care in the intervention group, and nearly complete follow-up in both the intervention and usual care sites.

Our study has important policy implications. The WHO has endorsed the HEARTS package for hypertension control since 2018, and by the end of 2022 over 12 million patients from 32 countries had been treated according to the HEARTS approach.^5^ Yet, no study had yet rigorously tested its effectiveness compared with usual hypertension care. To our knowledge, the Bangladesh HEARTS trial is the first to formally evaluated the WHO HEARTS package compared with usual care. Additionally, the results of this study support Bangladesh’s plan to offer the WHO HEARTS package in all of Bangladesh’s 492 UHC’s. Our results are also potentially generalizable to rural medical clinics in other countries with a similar system of health care delivery. Future studies should evaluate the HEARTS hypertension control package in different health care and health insurance contexts, in urban vs. rural areas, and in community-based care settings vs. standard clinics, with robust implementation evaluation. In rural Bangladesh, persons typically seek medical care in one of approximately 14,000 community clinics, as opposed to the nearest UHC, which is sometimes located at a considerable distance from patients’ homes. Scaling up patient-centered hypertension care in Bangladesh will require additional system changes to provide hypertension care and potentially care for other NCDs in these community clinics, which to date focus mostly on maternal and child healthcare.

In conclusion, in rural Bangladesh, the WHO-HEARTS package significantly lowered BP and improved hypertension control. These results provide evidence to scale up the WHO-HEARTS hypertension control package in Bangladesh and support its implementation in other low and middle-income countries.

## Data Availability

A complete, de-identified dataset representing the Bangladesh HEARTS trial is held at the trial coordinating center at Johns Hopkins University. Interested researchers may request access to this dataset.

## Acknowledgments

The authors thank the hypertension patients who participated in the HEARTS program evaluation, and we thank the dedicated health workers who made the HEARTS program and this study possible. We also thank the Medical Officers and Project Officers of the Bangladesh Hypertension Control Initiative (Bangladesh HEARTS) for their extraordinary efforts to complete this study and to deliver the HEARTS package of hypertension care to the people of Bangladesh living with hypertension. Medical Officers: Dr. Deb Dulal Dey Parag, Dr. Mahfuja Luna, Dr. Md Abbas Ibn Karim, Dr. Md. Ruhul Amin, and Dr. Md. Shahinul Islam. Project Officers: Dr. Iqbal Ahmed Khan, Khondokar Ehsanul Amin, Md. Riasat Razi Ullah, and Md. Sazzad Hossain.

